# Altered functional connectivity between cortical premotor areas and the spinal cord in chronic stroke

**DOI:** 10.1101/2024.04.08.24305494

**Authors:** Hanna Braaß, Silke Wolf, Jan Feldheim, Ying Chu, Alexandra Tinnermann, Jürgen Finsterbusch, Christian Büchel, Christian Gerloff, Robert Schulz

**Author notes:** Corresponding author: Hanna Braaß. These authors contributed equally to this work (Christian Gerloff, Robert Schulz).

## Abstract

**Background:** Neuroscience research has contributed significantly to understanding alterations in brain structure and function after ischemic stroke. Technical limitations have excluded the spinal cord from imaging-based research. Available data are restricted to a few microstructural analyses, and functional connectivity data are absent. The present study attempted to close this knowledge gap and assess alterations in corticospinal activation and coupling changes in chronic stroke.

**Methods:** Thirteen well-recovered stroke patients underwent corticospinal functional MRI while performing a simple force generation task. Task-related activation was localized in the ipsilesional primary motor cortex (M1), ventral premotor cortex (PMV), and supplementary motor area (SMA), as well as in the cervical spinal cord. Psycho-physiological interactions and linear modeling were used to infer functional connectivity between cortical motor regions and the cervical spinal cord and their associations with motor deficits.

**Results:** The main finding was that PMV and SMA showed topographically distinct alterations in their connectivity with the spinal cord. Specifically, we found a reduced coupling between SMA and the ipsilateral ventral spinal cord and an enhanced coupling between PMV and ventral and intermediate central spinal zones. Lower SMA- and higher PMV-related spinal cord couplings were correlated with residual deficits.

**Conclusion:** This work provides first-in-human functional insights into stroke-related alterations in the functional connectivity between cortical motor areas and the spinal cord, suggesting that different premotor areas and spinal neuronal assemblies might be involved in coupling changes. It adds a novel, promising approach to better understanding stroke recovery and developing innovative models to comprehend treatment strategies with spinal cord stimulation.

## Introduction

Imaging-based systems neuroscience research has significantly contributed to our current understanding of alterations in brain structure and function after ischemic stroke and how they relate to deficits and recovery processes. Apart from cognitive and language functions, deficits in the motor domain, particularly affecting the upper limb, are important sequelae after stroke, impeding private and professional rehabilitation^1^. Structural^2^ and functional imaging^3,4^ have built a robust body of evidence showing how acute stroke lesions affect brain activation and multi-site communication patterns, as well as the structural integrity of the human motor network and its key areas and interconnecting pathways comprising the primary motor cortices (M1), frontal secondary motor areas including the ventral premotor cortex (PMV), the supplementary motor area (SMA), subcortical brain regions, and the cerebellum. The corticospinal tract (CST), the motor network’s central outflow system, has been chiefly addressed structurally at various cerebral levels via lesion load^5^, fiber count^6,7^, and diffusion-imaging-based analyses of white matter microstructure^2,8^. Recently, advanced structural MRI extended the field of view towards the brain stem and the cervical spinal cord regarding structural CST alterations^9^. Such CST data were collectively reported to be critically linked to stroke recovery. Notably, studies showed that not only CST fibers originating from M1 but also secondary CST originating from non-primary motor areas, such as PMV and SMA^10^, show structure-outcome associations^8,11–15^. It has been repeatedly discussed that such CST might mediate corticospinal neurotransmission while bypassing lesioned CST components^16–20^. For instance, higher SMA activity in EEG^21^ and stronger structure-outcome associations for M1-PMV pathways^17,22^ were found mainly in patients with more CST damage. It has been speculated that these alternate CST trajectories might act on lower motor neurons directly or, more likely, indirectly via modulation of spinal interneuronal assemblies and circuits^23,24^. After earlier reports on the spinal cord and circuit dysfunctions after stroke^25^ and the first report of successful epidural cervical spinal cord stimulation for improvements of chronic post-stroke upper-limb paresis just recently^26^, the interest in the spinal cord after cortical stroke experienced a remarkable renaissance^27–29^. Animal data had already suggested that the spinal cord might be involved in recovery after cortical motor stroke^30–33^. However, functional data on spinal cord activations and functional connectivity with cortical motor networks are unavailable. Such analyses would be of great value to better understand stroke recovery in general and shed light on such groundbreaking innovative treatment protocols with spinal cord stimulation, in particular.

Advancements in functional MRI (fMRI) of the spinal cord^34,35^ and simultaneous cortical and spinal cord fMRI^36,37^ opened a new window to investigate functional corticospinal networks in humans with high spatial resolution^38^. In the motor domain, studies have just started to explore corticospinal functional connectivity during finger or hand movements in healthy participants^35,39,40^. In our previous study, we could link ventral spinal activation during simple hand movements not only to M1 activation but also to PMV activation^40^. This added first fMRI data to sparse transcranial magnetic stimulation data^41,42^, which suggested that human premotor areas might be functionally connected to the lower cervical spinal cord, likely interneurons, and contribute to distal upper limb functions in humans.

The present study was designed to explore alterations in cortical and spinal cord activation and changes in corticospinal coupling during a simple visually guided force generation task for the first time in human stroke survivors. Task-related activation was localized in ipsilesional PMV, and SMA, and the cervical spinal cord. Task-related spinal cord activation and psycho-physiological interactions (PPI) were inferred to assess functional connectivity between cortical motor regions and the cervical spinal cord. Linear modeling was used to address their associations with motor deficits.

## Materials and Methods

### Subjects

13 stroke patients and 13 age-matched healthy controls without any neurological damage unrelated to healthy aging were included in the analysis. Participants were right-handed and provided informed consent following the Declaration of Helsinki. The study was approved by the local ethics committee of the Medical Association of Hamburg (PV6026). See the Supplemental Material for more details.

### Motor Task

A simple motor task was used comprising repetitive whole-hand grips in a block design, as previously introduced in detail^40^. The stroke patients performed repetitive, visually guided, almost isometric whole-hand grips with their affected hand with three varying predefined force levels. The healthy controls performed the task with the right or left hand corresponding to the affected side of the matched patients (“pseudo-side”). See the Supplemental Material for details.

### Behavioral data

A JAMAR Hand Dynamometer (built by Patterson Medical, Warrenville, USA) measured both hands’ maximum whole-hand grip force. Further standardized tests included the National Institutes of Health Stroke Scale (NIHSS), the Fugl Meyer Assessment of the upper extremity (UEFM), the modified Rankin Scale (MRS), and the Nine-hole peg test (NHP). Healthy controls underwent grip force measurement and NHP testing. Relative NHP values were calculated by dividing the values of the affected and unaffected sides, or pseudo-sides.

### MRI data acquisition

A 3T Prisma MRI scanner (Siemens Healthineers, Erlangen, Germany) and a 64-channel combined head-neck coil were used to acquire cerebral and spinal imaging data. The MRI protocol was identical to the previously described MRI protocol^40^. Detailed sequence parameters are described in the Supplementary Material.

MRI data preprocessing and first-, and second-level analyses are similar to our previously described methods^40^. Detailed information is available in the Supplementary Material.

### Psycho-physiological interaction

Psycho-physiological interaction (PPI) models were implemented with seed regions in ipsilesional PMV and SMA to assess the functional connectivity between the brain and the spinal cord. The time course was extracted from spherical regions of interest (ROI) with a radius of 2mm around the individual peak voxel for each area in each subject. PPI models were calculated in the spinal cord with the extracted time course of each brain region. See the Supplemental Material for details.

### Further statistical analyses

The statistical package R 4.3.1^43^ was used for statistical analysis. Linear models were used to analyze age, NHP ratio, and maximum grip force for group-specific differences, with “side” as an additional confound of no interest. Two-sided t-tests were used to compare the exerted forces between the groups.

## Results

### Demographic, clinical and task data

13 chronic stroke patients (12 males and 1 female, all right-handed, aged 62.6±9.7 years, mean±SD) and 13 healthy controls (eight males and five females, all right-handed, aged 64.5±11.9 years) were included in this work. Clinical characteristics are given in **Tab. S*2***. There were no significant group differences for age (*P=0*.*65*) or maximum grip force (*P=0*.*75*). The NHP ratio was numerically higher among stroke patients (*P=0*.*056)*. A topographic map of the distribution of stroke lesions is shown in **Fig. 1**. During the motor task, the exerted target forces for stroke patients were 42.0±8.5%, 61.0±8.4%, and 76.7±10.97% for low, medium, and high levels, respectively. For controls, they were 43.7±12.2%, 60.2±8.0%, and 77.9±6.5%, respectively, with no group differences.

**Figure 1.**
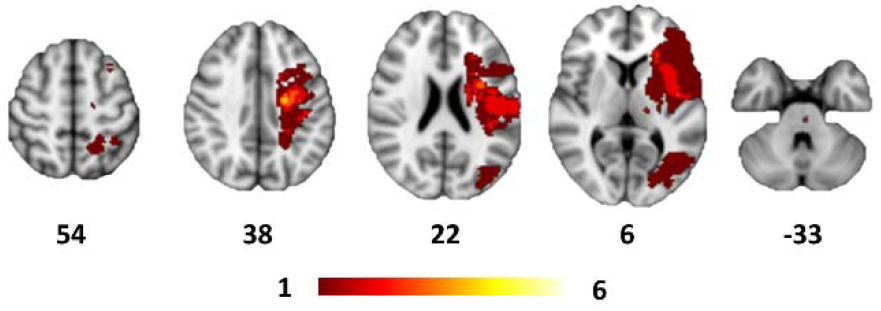
Topography of stroke lesions. All masks of stroke lesions are projected on the left hemisphere, overlaying a T1-weighted template in MNI space (z-coordinates below each slice). The color intensity indicates the number of subjects whose lesion voxels lie within the colored region.

### Spinal cord and cortical activation during force generation

Force generation across all three force levels led to a significant activation on the group level, primarily in the ipsilesional (= ipsilateral to the measured/affected hand) spinal cord between the lower parts of the C6 and C7 vertebral level, corresponding to the C7 and C8 spinal segments in both groups (**Fig. 2, A, B**). The stroke patients exhibited a higher activity controls, localized in the middle of the spinal than cord cross-section between vertebra C6 and C7 (**Fig. 2, C**). The cluster characteristics are listed in **Tab. S3**. In stroke patients, increasing force levels led to an increase in the spatial extent of BOLD responses from a focal activation at the C7 vertebral level towards more distributed spinal activations between C5 and C7 (**Fig. S2, A)**. In the control group, a similar increase was observed from a focal activation at the C5 vertebral level during low forces towards more distributed spinal activations between C5 and C7 (**Fig. S2, B**).

**Figure 2.**
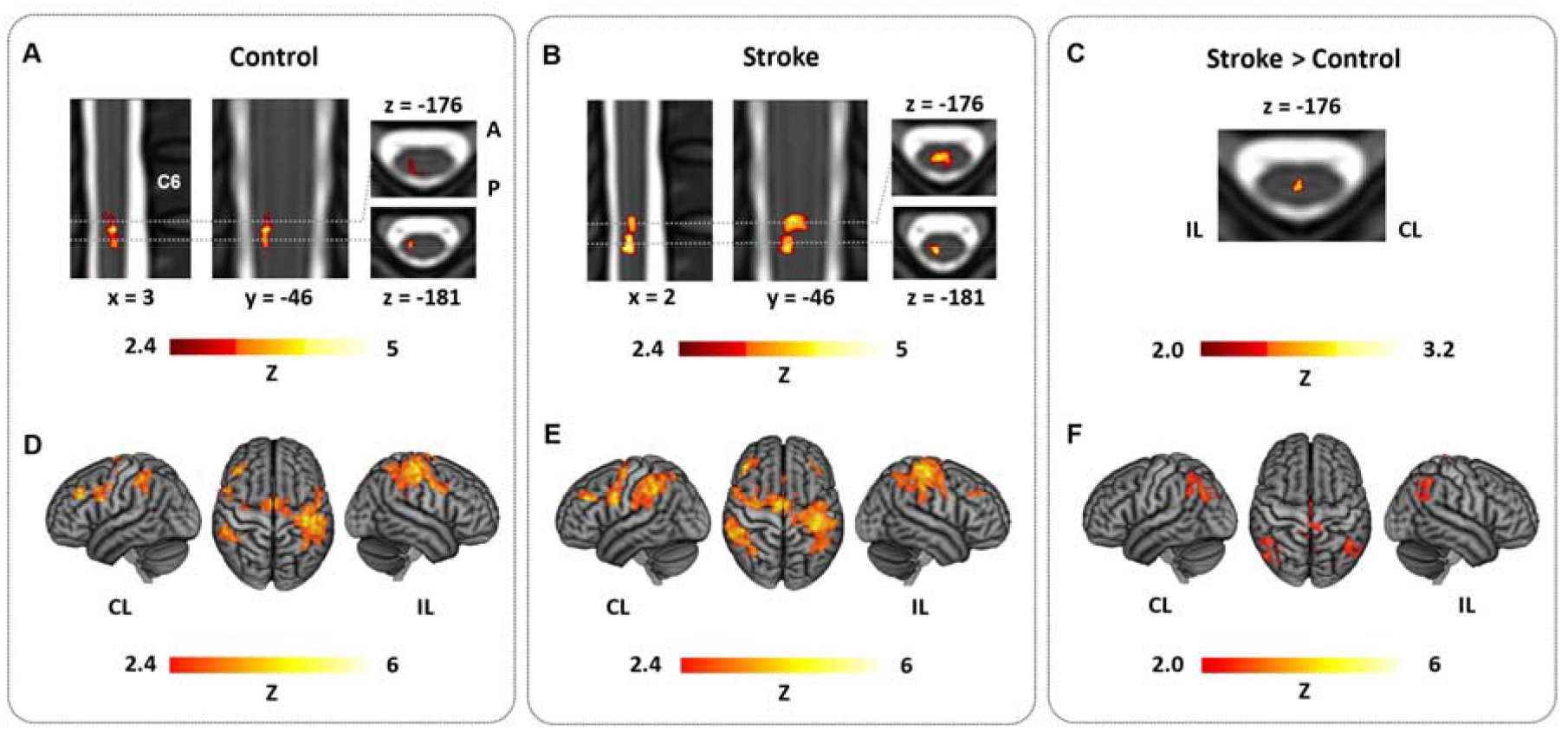
Topography of spinal cord and cortical brain activation during force generation. (**A**), (**B**): Group mean spinal activation during the task with maximum grip force, side, and age as additional confound parameters, (**A**): control group, (**B**): stroke group (**B**); (**C**) group comparison [stroke > control], controlled for maximum grip force; (**D**), (**E**): Group mean activation during the task with max. grip force, side, and age as additional confound parameters, (**D**): control group, (**E**): stroke group, (**F**) group comparison [stroke > control] controlled for max. grip force. Z-maps were thresholded by Z>2.4 (group mean) and Z>2.0 (group comparison), with a cluster significance threshold of *P*<0.05; Spinal cord activations are overlaid on the PAM50_t2-template. Cerebral activations are rendered on a T1 template in MNI space. IL = ipsilesional, CL = contralesional, A = anterior, P = posterior

Cerebral activity on group level was detected in both groups across force levels primarily in the ipsilesional primary sensorimotor cortex comprising M1 and the primary sensory cortex S1, in bilateral SMA, bilateral dorsal premotor cortex, bilateral PMV and a widespread activation in posterior parietal cortices along the intraparietal sulcus (**Fig. 2, D, E**). Group comparisons revealed higher SMA and bilateral intraparietal sulcus activity in the stroke group (**Fig. 2, F**). The results of the force-level specific analyses are shown in **Fig. S3**. Cluster characteristics of the task activation are listed in **Tab. S4**.

### Corticospinal connectivity during force generation

Psycho-physiological interactions (PPI) were computed to investigate task-specific functional corticospinal coupling. Compared to controls, the connectivity was enhanced between ipsilesional PMV and an intermediate spinal zone at C5 and ventral areas at the upper C6 vertebral levels (**Fig. 3, B**). The correlation analysis between functional connectivity and motor impairment revealed one cluster in the ventral spinal cord at C6-level, in which an increa**se** in connectivity was correlated with lower UEFM scores and, therefore, more severe motor impairment (**Fig. 3, C**). For fine motor skills, assessed via NHP ratio, one additional cluster was found in the area of the intermediate zone, also at the C6 level (**Fig. 3, C**). A sensitivity analysis excluding the three most severely affected stroke patients (UEFM<55) supports the results with higher connectivity in the stroke patients in the intermediate zone at C5 and an increasing **a**nd even positive coupling strength in patients with worse fine motor skills, i.e., higher NHP-ratios, at C6 (**Fig. S4**). Cluster characteristics are listed in **Tab. S5**.

**Figure 3.**
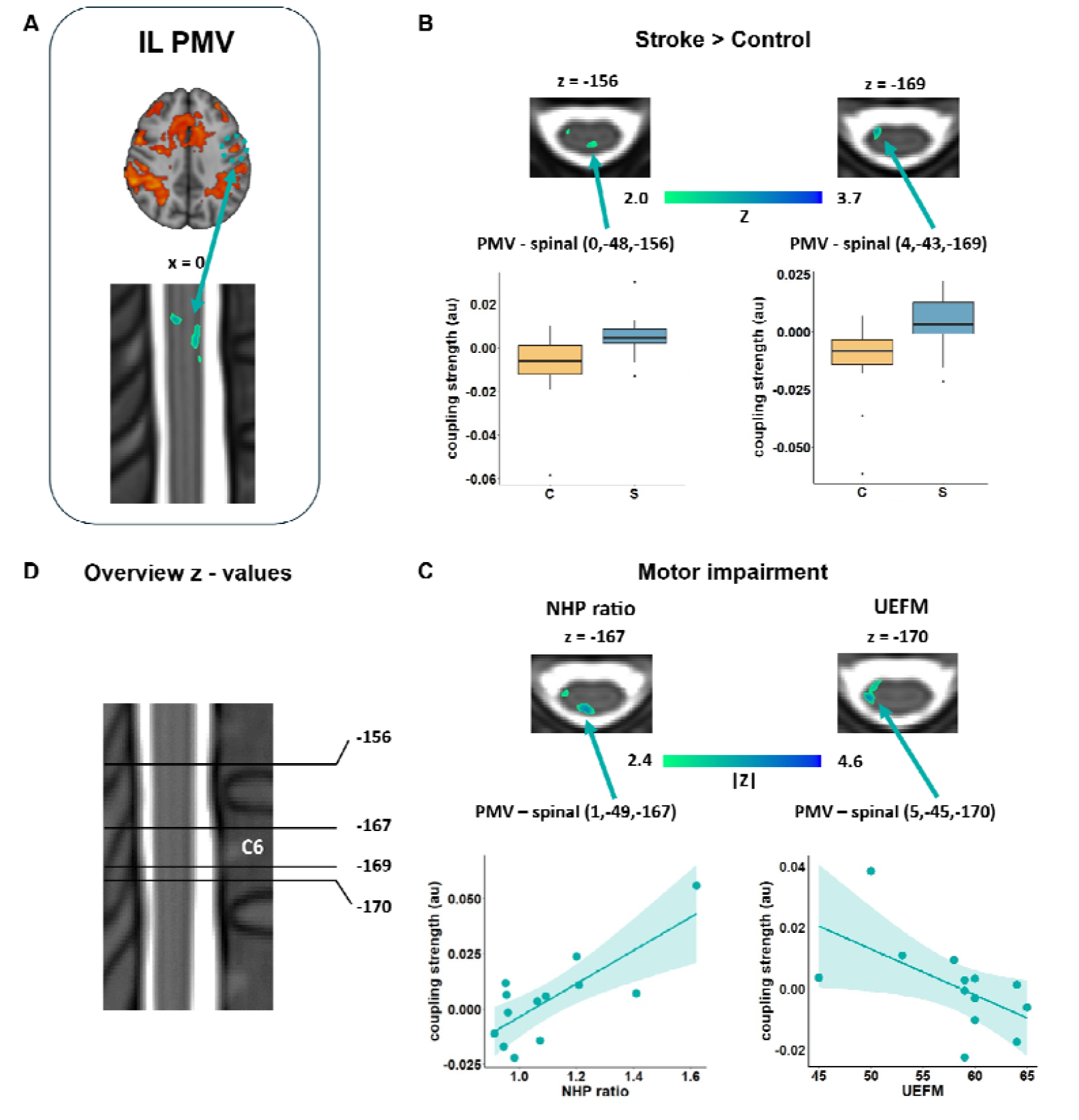
Alterations in corticospinal coupling during force generation for PMV. PPI analysis between ipsilesional PMV (**A**) and the spinal cord. (**B)** Group comparison Stroke > Control, controlled for maximum grip force, (**C)** Stroke patients: Correlation with motor impairment, controlled for maximum grip force, age, side, (**D)** Overview of the slices displ yed along the z-axis; (Z-maps are thresholded by Z>2.0 (group comparison) and |Z|>2.4 (Correlation analysis), cluster significance threshold of *P<0*.*05*). Spinal cord activations are overlaid on the PAM50_t2-template, and MNI coordinates are given. All images are in radiological orientation. The coupling strengths in spinal cluster voxels (MNI coordinates as indicated) were extracted and displayed for group comparison by boxplots and for correlation as a function of NHP ratio and UEFM. C = control, S = stroke.

For SMA, stroke patients exhibited a significantly reduced corticospinal connectivity between comprising ventral horn areas and surrounding white matter at ipsilesional and contralesional lower C6 vertebral level (**Fig. 4, B**). In the correlation analysis with motor impairment, we detected an ipsilesional ventral location in which reduced, i.e., negative connectivity, was correlated with more severe deficits. (**Fig. 4, C**). The sensitivity analysis without the three most severely affected stroke patients supports the results with lower connectivity in the patients i**n t**he ventral horn area at C6 (**Fig. S5**). The correlation analysis with NHP-ratio and UEFM showed no significant relationship in the described regions without these severely affected patients. Additional cluster results are listed in **Tab. S6**.

**Figure 4.**
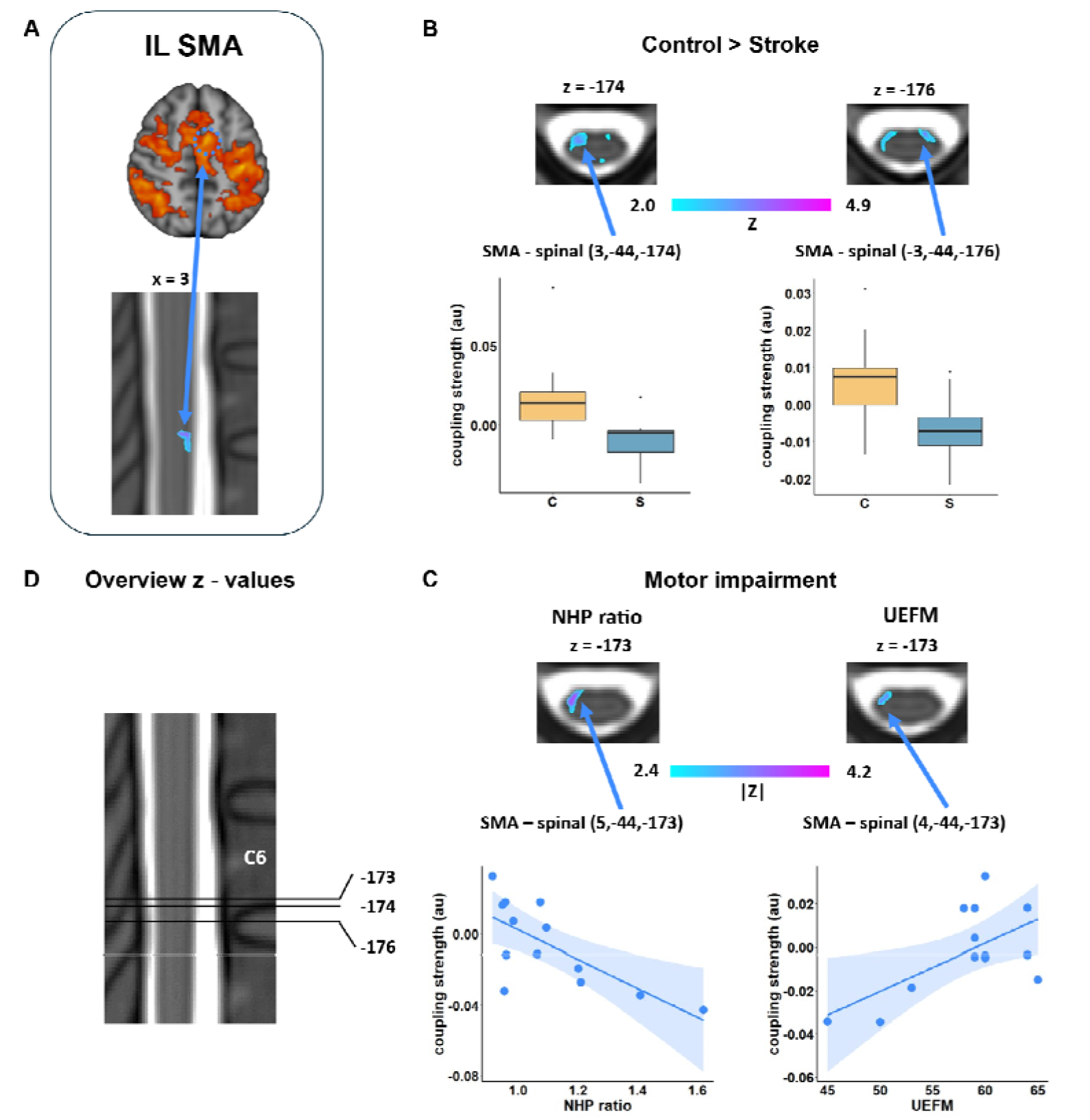
Alterations in corticospinal coupling during force generation for SMA. PPI analysis between ipsilesional SMA (**A**) and the spinal cord. (**B)** Comparison Stroke > Control, controlled for maximum grip force (**C)** Stroke patients: Correlation with motor impairment, controlled for maximum grip force, age, side, (**D)** Overview of the slices displayed along the z-axis; Z thresholds and image presentations are identical to **Figure 3**.

## Discussion

The main finding of the present study was that chronic stroke patients show alterations in activation and connectivity between cortical motor areas and the spinal cord while performing a simple force generation task with the affected hand. Concerning connectivity, topographically different and clinically relevant changes were found, which comprised the connectivity between PMV, SMA, and the spinal cord. They were opposite in the group comparison: for SMA, we found a reduced coupling with voxels localizing in the ventral spinal cord at lower C6 vertebral levels. For PMV, an enhanced coupling was detected with ventral regions at upper C6 vertebral levels and intermediate central spinal zones at C5-level. Finally, lower SMA- and higher PMV-related spinal cord coupling located at the C6-level, most likely contributing to the ipsilateral ventral areas, were directly correlated with persistent motor deficits. These data provide first-in-human functional insights into possible corticospinal network alterations after stroke, adding a novel, promising functional perspective to the emerging field of spinal cord analyses and stimulation in stroke recovery research.

The present work is based on previous animal and human imaging and electrophysiology data, arguing that the spinal cord may be considered an essential, however largely neglected, factor in modulating recovery after cortical stroke. For instance, previous animal work has evidenced an upregulation of spinal structural plasticity, neurotrophins, and cytokines^44,45^. Recent transcriptome analyses found gene upregulations related to neurite outgrowth in stroke-denervated spinal gray matter, particularly in its intermediate laminae^30^. One study reported spinal axonal sprouting and microglial activation to support presynaptic site formation^31^, potentially occurring in central spinal regions where axonal outgrowth has been consistently documented in stroke animal models^32,46^. Interestingly, at the group level, we also found increased spinal cord activation in similar central voxels. Furthermore, stronger PMV coupling in central voxels was correlated with greater deficits. Speculative in nature, such areas might correspond to intermediate zones such as lamina X or, even more specifically, interneurons from V0 families, which were reported to contribute to complex corticospinal circuits connecting motor and sensory cortical influences with spinal neuronal assemblies^23,24^. Animal studies suggest that these assemblies are involved, for example, in left-right coordination^47^ or aberrant excitation during the progression of amyotrophic lateral sclerosis^48^.

As a second topographically interesting finding, alterations in corticospinal coupling were detected for areas localizing towards the ipsilateral ventral spinal cord. Particularly for SMA, the overlap between voxels exhibiting reduced connectivity and voxels exhibiting a correlation with behavior was notable. Ipsilateral ventral horn areas would align with the location of lower motor neurons or V1 interneuron populations comprising multiple cell types, including Renshaw cells and Ia inhibitory interneurons^49–51^. It remains elusive if any, or which neuronal component might be mainly related to the associations in the present fMRI datasets. At least from a very simplified clinical view, the patients included in this analysis did not show hand spasticity in hand/finger flexion and extension. Larger samples with varying degrees of spasticity could allow for regression modeling of this valuable covariate. Combined approaches with electrophysiology, such single-fiber electromyography, could add to the regression of trans-synaptic degeneration of spinal motor neurons after stroke^52^.

Recent human data have convergingly evidenced, potentially against previous assumptions derived from non-human primates^53–56^, that PMV and SMA might be structurally^57^ and functionally connected^41,58^ not only with upper segments of the spinal cord but also with lower cervical spinal cord contributing to distal upper limb functions. These connections will likely be maintained by poly-synaptic connections along spinal interneural routes, including the propriospinal system^59^. The present data builds on this knowledge. They confirm that premotor areas may be functionally connected to the lower spinal cord and provide evidence that post-stroke connectivity is subject to changes associated with persistent clinical deficits. Unfortunately, we can only speculate about the meaning of the directions of alterations and linear associations. For instance, the negative coupling between SMA and the ventral spinal cord could be interpreted as an inhibitory process during the execution of the task^60^. How this coupling might mechanistically explain impairment remains open for discussion and controversy^3,4,21,61^.

Functional analyses of corticospinal coupling and spinal cord activation might help to better understand stroke recovery in general. However, this approach might be promising to comprehend the effects of spinal cord stimulation after stroke, especially in light of recent breakthrough results demonstrating improvements in arm and hand paresis in two chronic stroke patients^26^. It has been speculated that spinal cord stimulation might act via the recruitment of primary afferents, which provide excitatory input to motoneurons and interneurons directly connected to such afferents. Thereby, spinal stimulation might increase the responsiveness to residual cortical inputs, e.g., from preserved primary and non-primary CST or alternate descending pathways, including the reticulospinal tract^2,9,62,63^. Enabling researchers to assess changes in corticospinal activation and connectivity with sufficient spatial resolution, as illustrated in our study, corticospinal fMRI could contribute to disentangling mechanisms such as increases in motoneuron excitability, postsynaptic inhibition or sensory gating, which have been discussed as potential mechanisms for spinal cord stimulation^27^.

It can be argued that the most severely affected patients significantly influenced the analysis results. For this reason, we performed a sensitivity analysis in which patients with UEFM<55 were excluded. However, this analysis showed that the group comparison results in the ventral areas for SMA and especially PMV remained stable. The correlation for PMV in the intermediate zone was also stable, indicating that the connectivity changes for SMA occur more clearly in the severely affected patients. However, changes for PMV, in particular, also occur in less severely affected patients.

There are several other significant limitations to note. First, the small sample size must be considered when interpreting the results. Despite technical advances, spinal cord imaging remains a time-consuming challenge, particularly for older participants. Inclusion criteria were rather strict; more severely impaired patients could not perform the task correctly and could not participate in this study. Future studies on larger sample sizes are needed to verify the present results and may help to generalize them to the broader population. Second, the design of this study was cross-sectional. A longitudinal approach could provide further insights into the temporal dynamics of stroke-related corticospinal coupling and activation changes. Thirdly, due to the technically limited resolution, smoothing, and structure of the spinal cord, the spatial localization of the activated spinal regions can only be indicated approximately; further studies will be necessary to verify the results presented here. Congruent with our previous work in younger, healthy participants^40^ and given broad evidence suggesting that PMV and SMA are key areas that undergo stroke-related changes in brain structure and function^2–4^, we focused our analyses on these cortical areas.

Subcortical nodes of the human motor network, such as the basal ganglia, the cerebellum, and the brainstem, could not be included as seed areas in this work because of the reduced fields of view due to technical restrictions. Finally, the spinal resolution was limited, complicating the inference of precise topographical interpretations regarding spinal neuronal assemblies.

## Conclusion

Collectively, this work provides first-in-human insights into stroke-related alterations in the functional connectivity between cortical motor areas and the spinal cord. The analysis of the results suggests that different premotor areas and spinal neuronal assemblies might be differentially prone to and involved in coupling changes. This study adds a novel, promising approach to better understanding stroke recovery in general and developing innovative models to comprehend groundbreaking treatment strategies with spinal cord.

## Data Availability

All data needed to evaluate the conclusions in the paper are present in the paper and/or the Supplementary Materials. Processed brain and spinal cord activation data and demographic data to reproduce the findings are available from the corresponding author upon reasonable request.

## Author Contributions

Conceptualization: HB, CG

Investigation: HB, JFE, SW

Methodology: JFE, HB, JFI, YC, CB, AT

Formal Analysis: HB, RS

Writing – Original Draft Preparation: HB, RS

Writing – Review & Editing: HB, RS, AT, CB, CG, JFI, YC, SW

