## Supplementary Material for "Altered functional connectivity between cortical premotor areas and the spinal cord in chronic stroke"

### Supplemental Methods

#### Subjects

Fourteen chronic stroke patients participated in the study. Inclusion criteria were first-ever ischemic stroke ≥ 6 months with a persistent motor deficit involving the hand function and age between 18 and 80 years. Exclusion criteria were preexisting clinically silent brain lesions > 1cm$^3$ on MRI, pre-existing motor deficits, contraindications against MRI, psychiatric disease, and active drug abuse. One patient had to be excluded from the final analysis because he terminated the MRI experiment prematurely. The resulting 13 patients were matched with 13 age-matched healthy controls without any neurological damage unrelated to healthy aging. Participants were right-handed and provided informed consent following the Declaration of Helsinki. The study was approved by the local ethics committee of the Medical Association of Hamburg (PV6026).

#### Motor Task

A simple motor task was used comprising repetitive whole-hand grips in a block design, as previously introduced in detail[1]. In three active conditions, stroke patients performed repetitive, visually guided, almost isometric whole-hand grips with their affected hand with three varying predefined force levels (low, medium, and high corresponding to 30%, 50%, and 70% of the maximum output measurement) using an MRI compatible grip force response device (Grip Force Bimanual, Current Design, Inc, Philadelphia, PA). Healthy controls performed the task with the right or left hand corresponding to the affected side of the matched patients ("pseudo-side"). In the active condition, a white cross on a screen blinked at 0.8Hz, indicating the frequency of hand grips. Details of the time course of each block have been reported previously[1].

#### MRI data acquisition

A 3T Prisma MRI scanner (Siemens Healthineers, Erlangen, Germany) and a 64-channel combined head-neck coil were used to acquire cerebral and spinal imaging data. The MRI protocol was identical to the previously described MRI protocol[1]. The imaging modalities included high-resolution T1-weighted cerebral and spinal cord, T2*-weighted spinal cord, and task-related fMRI cerebral and spinal cord images with the iso-center approximately centered to vertebral level C2/C3. For the T1-weighted sequence, a 3-dimensional magnetization-prepared rapid gradient echo (3D-MPRAGE) sequence was used, which covered the head, the cervical spine, and the upper part of the thoracic spine with the following parameters: repetition time (TR) = 2300ms, echo time (TE) = 3.4ms, flip angle 9°, 236 coronal and 320 axial slices, voxel size: 1.0 x 1.0 x 1.0mm$^3$. The T2*-weighted image (MEDIC sequence) covered the identical part of the cervical spine as the functional spinal slices, centered on vertebra C6, with the following parameters: TR = 307ms, TE = 21ms, flip angle 20°, eight axial slices, voxel size: 0.5 x 0.5 x 5.0mm$^3$. For task-related fMRI, a combined corticospinal protocol based on echo-planar imaging (EPI) was used to record BOLD responses in the brain and spinal cord [2,3] covering 32 slices, divided into two sub-volumes (**Fig. S1**). These two sub-volumes had different geometry, timing parameters, and shim settings [4]. The shim settings were determined using a field map acquisition and a dedicated shim algorithm [4]. The upper volume included 24 axial slices (voxel-size: 2.0 x 2.0 x 2.0mm$^3$, 1mm gap between slices) in the brain. The lower sub-volume consisted of 8 axial slices (voxel-size: 1.0 x 1.0 x 5.0mm$^3$, no gap between slices), centered at the vertebral body of C6 and covered the vertebral bodies of C5, C6, and C7. The whole sequence was measured with the following parameters: TR = 2231ms, TE = 30ms/31ms (brain/spinal cord), flip angle = 75°. Additionally, one whole-brain EPI volume was measured with the following parameters: TR = 2385ms, TE = 30ms, flip angle 75°, 36 axial slices, voxel size: 2.0 x 2.0 x 2.0mm$^3$, 1mm gap between the slices. During the fMRI sessions, pulse, respiration, and the trigger signal were recorded (sampling rate = 400Hz) using the Physlog-function (Ideacmdtool) and respiratory and pulse measurement devices provided by Siemens Healthineers, Erlangen, Germany.

**Image preprocessing**

In line with our previous study[1], brain and spinal cord images were pre-processed separately. The brain fMRI images were pre-processed using the Oxford Center for fMRI of the Brain's (FMRIB) Software Library (FSL) v. 6.0.4 [5]. Un-flipped data were used for the readout of the individual BOLD parameters. For the second-level analysis, all T1-weighted and EPI brain images with right-sided stroke lesions were flipped to the left hemisphere. This hemispheric flip was also performed in the matched controls. The whole-brain EPI image of each subject was linearly co-registered to the individual high-resolution T1-weighted brain images, and the individual T1-weighted image was linearly co-registered with the MNI152 T1-2mm image from the FSL library. The transformation matrices were concatenated for further preprocessing steps. The first five dummy volumes of the task-related fMRI images were discarded, and the averaged fMRI image was registered to the whole brain EPI image. The concatenated transformation matrices were then used for registration to the MNI152-T1-2mm image. The fMRI images were further pre-processed with motion correction using MCFLIRT[6], and the images from both sessions were concatenated into one time series at the subject level.

The spinal fMRI images were pre-processed using the Spinal Cord Toolbox, v. 5.2[7], and FSL v. 6.0.4[5]. For the second level analysis, all T2*-weighted and spinal EPI images of patients with right-sided stroke lesions and their matched controls were flipped to the right side of the spinal cord and the un-flipped data were used for readout of individual BOLD parameters. The first step was that the spinal fMRI images were cropped with the spinal cord at the center of the image. Motion correction was performed using two phases of movement correction: MCFLIRT[6] was used for the first phase with spline interpolation and a normalized correlation cost function. The images across the two runs were realigned to the first image of the first run with a three-dimensional rigid-body realignment. To correct slice-independent motion, the second phase of motion correction was performed with two-dimensional rigid realignment independently for each axial slice[8,9]. The images from both sessions were concatenated into one time series at the subject level.

The spatial normalization from native to MNI standard space was performed using tools from the open-source Spinal Cord Toolbox [7,9]: The structural T2*-weighted images were normalized to the PAM50_T2s-template (resolution = 0.5 x 0.5 x 0.5mm³) [9,10]. After motion correction, the mean functional image was segmented to identify the spinal cord. The resulting binary spinal cord mask and the reversed deformation fields of the structural normalization were used to register the PAM50_T2-template on the mean functional image. The inverted resulting deformation field was then used for image normalization to PAM50-space. All normalized images were visually inspected for quality control at each step.

**Physiological noise modeling**

Cardiac and respiratory cycles are significant noise sources in spinal cord fMRI and can confound signal detection[9]. The SPM (SPM12) based PhysIO Toolbox version 8.1.0[11], ran in MATLAB version R2018a, was used to calculate the noise regressors. This toolbox uses a model-based physiological noise correction, which uses retrospective image correction (RETROICOR) of physiological motion effects[12], heart rate variability[13], and respiratory volume per time[14]. Based on the physiological signals, 18 noise regressors were generated. A cerebrospinal fluid (CSF) regressor was also generated from the CSF signal surrounding the spinal cord using a subject-specific CSF mask generated from the PAM50_csf-template[9,10].

**First level analyses**

Two different first- and second-level analyses were performed. The analyses of the cerebral and spinal images were conducted separately. For both analyses, the same explanatory variables (EVs) were used in the design matrices of the general linear models (GLM).

First, first-level analysis: For each volume (TR), the averaged maximum force produced was calculated and used as EV in the design matrix and, from now on, referred to as activation during the task. In the second first-level analysis, the low, medium, and high force levels were used as EVs. In both analyses, the temporally jittered instruction period[1] was separately modeled as an additional EV but not further analyzed in group analysis[15]. The brain and spinal cord data were analyzed separately.

Brain images: the motion-corrected functional images were spatially smoothed with a Gaussian kernel of 5mm full-width half maximum (FWHM) and high-pass filtered (90s) using the fMRI Expert Analysis TOOL (FEAT v6.00)[16,17]. Statistical maps of the pre-processed time series were generated using FMRIB's improved Linear Model (FILM) with pre-whitening[9,17]. The design matrices included the hemodynamic response function (gamma convolution, phase 0s, standard deviation 3s, mean lag 6s) convolved task vectors as EVs. Motion parameters and motion outliers, determined using *fsl_motion_outliers,* were entered as covariates to remove movement-related variance. The voxels of peak activation in the subject-specific activation maps for the task-related activation EV were localized in ipsilesional PMV, and SMA, according to the Harvard-Oxford atlas (SMA) and the HMAT template (Human motor area template)[18] (PMV) for correct localization (**Tab. S1**). For the second-level analyses, spatial normalization of the statistical images from the subject-level analyses to the MNI template was performed.

Spinal images: the motion-corrected functional images were spatially smoothed with a Gaussian kernel of 2 x 2 x 5mm FHWM, and the spinal cord was extracted from the data using a spinal cord mask, which was created from the PAM50_cord_template and spatial transformed in the subject-specific space. The data were further analyzed with FEAT and high pass filtered (90s). The statistical maps of the pre-processed time series were generated using FILM with pre-whitening. The design matrices included the hemodynamic response function (gamma convolution, phase 0s, standard deviation 3s, mean lag 6s) convolved task vectors as EVs, the physiological noise regressors, the CSF time series, the motion parameters and motion outliers as covariates of no interest. The voxels of peak activation in the subject-specific activation maps for the task-related activation were localized in the ipsilesional (ipsilateral to the moving hand) spinal cord. For the second-level group analysis, spatial normalization of the statistical images from the subject-level analyses to the PAM50-template was performed.

#### Psycho-physiological interaction

Psycho-physiological interaction (PPI) models were implemented with seed regions in ipsilesional PMV and SMA to assess the functional connectivity between the brain and the spinal cord. The time course was extracted from spherical regions of interest (ROI) with a radius of 2mm around the individual peak voxel for each area in each subject. PPI models were calculated in the spinal cord with the extracted time course of each brain region. The models contained the same task regressors as in the model of the whole task, the mean-centered time course, and the PPI interaction term with the task activation. In addition, the models contained all regression terms for movement and physiological noise as in the original GLM. Group comparison was calculated for each region with individual maximum grip force as an additional confound parameter using FLAME 1 and 2. The Z-maps were thresholded using a *Z-Score>2.0* with a cluster significance threshold of $P<0.05$. In two additional analyses in the stroke patients, demeaned relative NHP and demeaned UEFM were included to analyze the relationship between motor impairment after stroke and functional connectivity, corrected for maximum grip force, age, and side. The Z-maps were thresholded using a *Z-Score>2.4* with a cluster significance threshold of $P<0.05$. The estimated coupling strength was extracted from different spinal voxels (the specific coordinates are given in the figures showing the PPI results). To assess the robustness of the results, the PPI analyses were repeated without the three most severely impaired stroke patients (UEFM<55) and their matched controls.

#### Second level analyses

Each group's average activation maps were generated with the demeaned individual maximum grip force of the measured hand, side, and age as additional covariates using FMRIB's Local Analysis of Mixed-effects (FLAME) stages 1 and 2[16,19]. The group average activation maps were thresholded using a *Z-Score > 2.4* with a cluster significance threshold of $P<0.05$ to correct for multiple comparisons using GRF (Gaussian Random Field) theory[20]. Group comparison was calculated for the task activation with maximum grip force as an additional covariate. The activation maps for the contrasts [stroke > control] and [control > stroke] were thresholded using a *Z-Score>2.0* with a cluster significance threshold of $P<0.05$.

The estimated coupling strength was extracted from different spinal voxels (the specific coordinates are given in the figures showing the PPI results) and presented in a graphical manner.

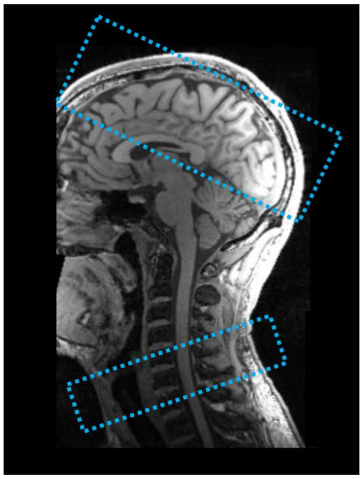

**Figure S1. Exemplary representation of the position of the two sub-volumes**

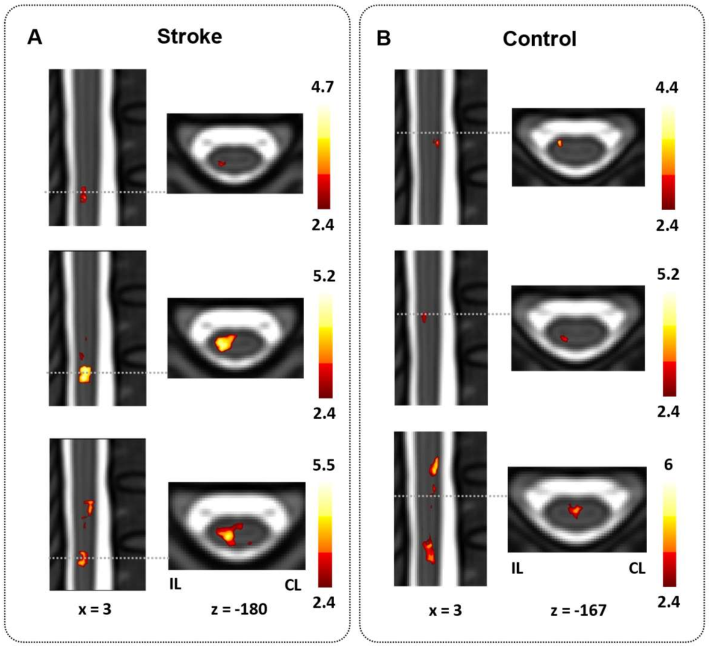


**Figure S2. Topography of spinal cord activation during force generation**
**(A, B)**: Estimated group mean spinal BOLD response (Z-maps, thresholded by *Z>2.4*, cluster significance threshold of *P*<0.05, maximum grip force, side, age were included as additional confound parameter) across the three force levels low, medium, high on one sagittal, (**A**)**:** stroke patients, (**B**)**:** control group; Spinal cord activations are overlaid on the PAM50_t2-template and are in radiological orientation.

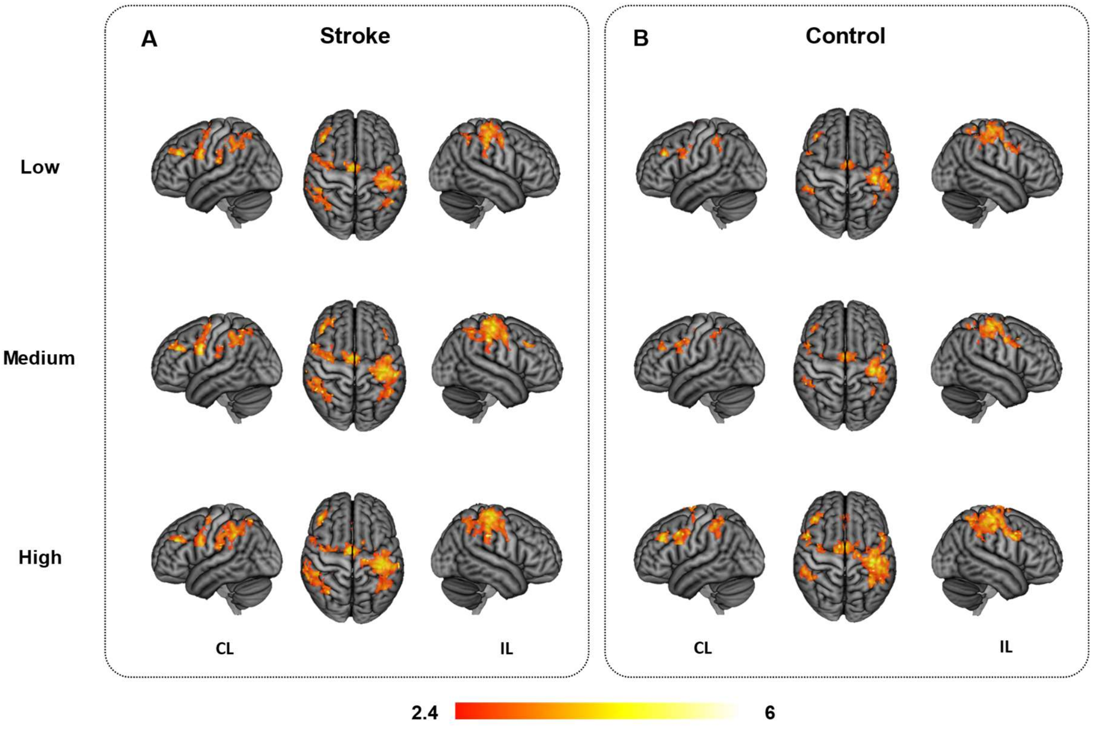


**Figure S3. Topography of cerebral activation during force generation**
**(A, B)**: Estimated group mean cerebral BOLD response (Z-maps, thresholded by $Z>2.4$, cluster significance threshold of $P<0.05$, maximum grip force, side and age were included as additional confound parameter) across the three force levels low, medium, high (**A**)**:** Stroke, (**B**)**:** healthy controls; Cerebral activations are overlaid on the T1 template in MNI space and are in radiological orientation.

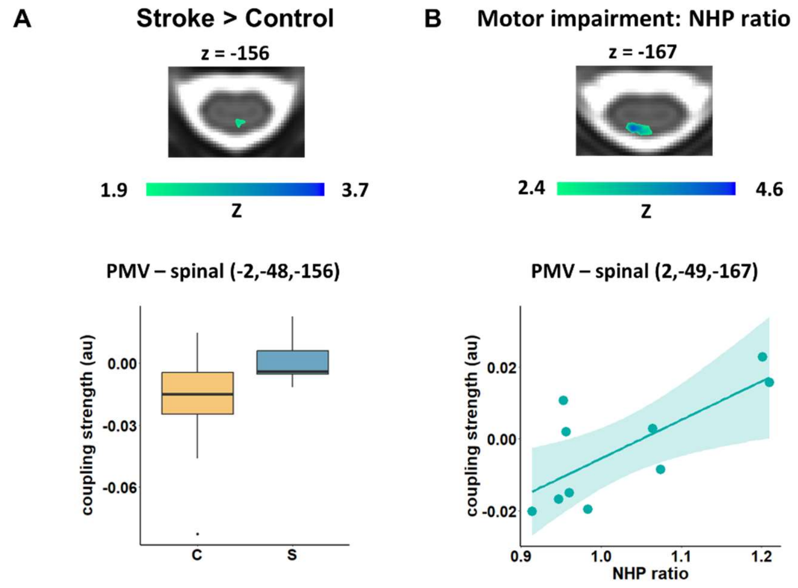


**Figure S4. PPI: IL PMV – spinal cord, sensitivity analysis**
PPI analysis between IL PMV and the spinal cord. **B** Group comparison Stroke > Control, controlled for maximum grip force, **C** Stroke patients: Correlation between PPI PMV - spinal and impairment of fine motor skills controlled for maximum grip force, age, side. (Z-maps are thresholded by $Z>1.9$ (group comparison) and $|Z|>2.4$ (Correlation analysis), cluster significance threshold of $P < 0.05$).

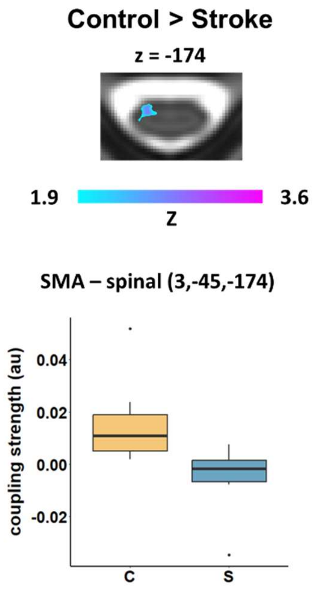


**Figure S5. PPI: IL SMA – spinal cord, sensitivity analysis**
PPI analysis between IL SMA and the spinal cord: Group comparison Control > Stroke, controlled for maximum grip force, **C** Stroke patients: (Z-maps are thresholded by Z>1.9 (group comparison), cluster significance threshold of P < 0.05).

**Table S1. Mean MNI coordinates of the individual peak voxel in IL SMA, PMV**

| Stroke | x | y | z | Control | x | y | z |
|---|---|---|---|---|---|---|---|
| **IL SMA** | -4 | -10 | 58 | **IL SMA** | -4 | -8 | 60 |
| **IL PMV** | -55 | 4 | 28 | **IL PMV** | -46 | 6 | 23 |

The peak voxel locations were projected on the left hemisphere (lesioned hemisphere, IL)

**Table S2. Characteristics of patients and healthy controls**

| ID | Age | Sex | Lesioned hemisphere | Lesion volume [ml] | Time since stroke [months] | Maximum Grip force [Kg] | UEFM | NHP ratio [aff/unaff] | MRS | NIHSS |
|---|---|---|---|---|---|---|---|---|---|---|
| S1 | 35-40 | m | Left | 0.1 | 43 | 49.7 | 65 | 0.95 | 1 | 1 |
| S2 | 61-65 | m | Left | 1.2 | 118 | 38.0 | 59 | 1.06 | 1 | 0 |
| S3 | 61-65 | m | Left | 0.4 | 12 | 30.0 | 60 | 0.91 | 1 | 0 |
| S4 | 71-75 | m | Right | 36.8 | 159 | 28.7 | 45 | 1.41 | 2 | 2 |
| S5 | 56-60 | m | Left | 0.3 | 9 | 39.3 | 64 | 0.96 | 1 | 0 |
| S6 | 66-70 | m | Left | 0.3 | 55 | 40.7 | 50 | 1.62 | 2 | 1 |
| S7 | 61-65 | m | Left | 0.1 | 56 | 48 | 60 | 1.20 | 1 | 1 |
| S8 | 66-70 | m | Right | 34.4 | 7 | 50.7 | 59 | 1.07 | 1 | 1 |
| S9 | 71-75 | w | Right | 3.4 | 6 | 25.3 | 59 | 1.21 | 1 | 0 |
| S10 | 56-60 | m | Left | 31.6 | 53 | 48.7 | 53 | 1.09 | 2 | 3 |
| S11 | 66-70 | m | Right | 64.9 | 16 | 39.0 | 58 | 0.95 | 1 | 0 |
| S12 | 46-50 | m | Left | 4.3 | 65 | 54.7 | 64 | 0.96 | 1 | 0 |
| S13 | 61-65 | m | Left | 0.7 | 20 | 40.7 | 60 | 0.98 | 1 | 0 |
| Patients | 62.6 (9.7) | 1 W / 12 M | 9 Left / 4 Right | 13.74 (21.08) | 47.62 (46.32) | 41 (9) | 58.15 (5.73) | 1.11 (0.21) | 1.23 (0.44) | 0.69 (0.95) |
| Controls | 64.5 (11.9) | 5 W / 8 M | 9 Left / 4 Right[a] | | | 40 (10)[b] | | 0.98 (0.13)[b] | | |

Characteristics of all patients individually and averaged per group for patients and controls. Mean values and standard deviation in brackets are given. Examination date of MRI imaging in months after stroke. UEFM = Upper Extremity Fugl Meyer Assessment. MRS = modified Rankin Scale. [a] healthy controls: examined side, left = right hand / left hemisphere, right = left hand / right hemisphere. [b] healthy controls: aff = examined hand.

**Table S3. FMRI-derived task-related spinal activation.**

| Contrast | x | y | z | Z | Cluster size |
|---|---|---|---|---|---|
| **Stroke** | 2.5 | -47 | -182 | 5.16 | 1290 |
| **Control** | 3.5 | -46 | -178 | 4.95 | 503 |
| **Stroke > Control** | 0 | -46 | -176 | 3.21 | 267 |

Group mean spinal BOLD activation: Stroke, $Z>2.4$; healthy control group, $Z>2.4$; and comparison Stroke > Control, $Z>2.0$; cluster significance threshold of $P<0.05$. Peak coordinates and cluster sizes are given.

**Table S4. FMRI derived task-related brain activation.**

| | Region | x | y | z | Z | Cluster Size |
|---|---|---|---|---|---|---|
| **Stroke** | IL Prec./Postc. G. | -40 | -22 | 52 | 5.57 | 13727 |
| | CL SMA | 6 | -4 | 58 | 5.3 | |
| | CL Prec. G. | 30 | -10 | 56 | 5.04 | |
| | CL Prec. G. | 52 | 4 | 28 | 5.03 | |
| | IL SMA | -2 | -10 | 56 | 4.98 | |
| | unspecified | 32 | -8 | 36 | 4.97 | |
| | CL Sup. Parietal L. | 38 | -50 | 52 | 5.03 | 4075 |
| | CL Supramarginal G. | 44 | -38 | 40 | 4.96 | |
| | CL Lateral Occ. C. | 30 | -64 | 40 | 4.94 | |
| | CL Supramarginal G. | 58 | -38 | 20 | 4.82 | |
| | CL Supramarginal G. | 58 | -42 | 40 | 4.81 | |
| | CL Sup. Parietal L. | 34 | -50 | 46 | 4.71 | |
| | CL MFG | 44 | 36 | 30 | 5.15 | 384 |
| | CL MFG | 44 | 28 | 30 | 4.11 | |
| | CL Frontal Pole | 36 | 46 | 30 | 3.21 | |
| | CL MFG | 40 | 30 | 40 | 2.96 | |
| | CL Frontal Pole | 30 | 44 | 36 | 2.92 | |
| | CL MFG | 50 | 24 | 26 | 2.84 | |
| **Control** | IL Prec./Postc. G. | -46 | -22 | 58 | 5.17 | 10546 |
| | IL Postc. G. | -46 | -30 | 48 | 5.15 | |
| | CL SMA | 4 | -2 | 56 | 5.06 | |
| | IL Prec./Postc. G. | -38 | -22 | 58 | 5.04 | |
| | IL Prec./Postc. G. | -36 | -28 | 62 | 4.92 | |
| | IL Sup. Parietal L. | -32 | -42 | 40 | 4.85 | |
| | CL AIPS | 28 | -50 | 32 | 4.64 | 1213 |
| | CL Supramarginal G. | 46 | -42 | 48 | 4.55 | |
| | CL Sup. Parietal L. | 40 | -46 | 52 | 4.35 | |
| | CL Supramarginal G. | 54 | -38 | 52 | 4.32 | |
| | CL Supramarginal G. | 52 | -32 | 42 | 4.1 | |
| | CL Supramarginal G. | 46 | -38 | 42 | 3.79 | |
| **Stroke > Control** | Prec. G./Cing. G. | 0 | -18 | 48 | 3.48 | 1773 |
| | Prec. G. | 0 | -26 | 56 | 3.31 | |
| | CL Prec./Postc. G. | 2 | -30 | 56 | 3.3 | |
| | IL SMA | -4 | -18 | 48 | 3.23 | |
| | IL SMA | 0 | -12 | 48 | 3.23 | |
| | IL Prec./Postc. G. | -6 | -32 | 72 | 3.2 | |
| | CL Lateral Occ. C. | 46 | -64 | 36 | 3.57 | 831 |
| | CL Supramarginal G. | 48 | -50 | 48 | 3.01 | |
| | CL Supramarginal G. | 56 | -50 | 34 | 3 | |
| | CL Lateral Occ. C. | 52 | -70 | 26 | 2.92 | |
| | CL Lateral Occ. C. | 46 | -74 | 30 | 2.79 | |
| | CL Lateral Occ. C. | 34 | -80 | 28 | 2.74 | |
| | IL Supramarginal G. | -44 | -54 | 48 | 3.26 | 570 |
| | IL Lateral Occ. C. | -50 | -66 | 32 | 2.82 | |
| | IL Lateral Occ. C. | -56 | -62 | 30 | 2.78 | |
| | IL AIPS | -38 | -60 | 40 | 2.63 | |
| | IL Lateral Occ. C. | -40 | -62 | 44 | 2.63 | |
| | IL Supramarginal G. | -60 | -56 | 30 | 2.61 | |

Group mean cerebral BOLD activation, Stroke, $Z>2.4$, Healthy control group, $Z>2.4$; comparison Stroke > Control, $Z>2.0$; IL=ipsilesional, CL=contralesional, G=Gyrus, C=Cortex, L=Lobule, SMA=Supplementary Motor Cortex, Prec. G=Precentral Gyrus, Postc. G=Postcentral Gyrus, AIPS= Anterior intra-parietal sulcus; cluster significance threshold of $P<0.05$.. Identification of the regions with the Harvard-Oxford atlas and Juelich Histological Atlas.

**Table S5. Cluster values of group comparison: PPI between cerebral seeds and spinal**

| PPI | comparison | x | y | z | Z | Cluster size |
|---|---|---|---|---|---|---|
| PPI IL SMA - spinal | Control > Stroke | -3 | -44 | -176 | 3.76 | 378 |
| | | -3 | -46 | -160 | 3.38 | 353 |
| | | 3 | -44 | -174 | 4.59 | 269 |
| PPI IL PMV - spinal | Stroke > Control | 0 | -48 | -156 | 3.71 | 809 |

Threshold *Z>2.0*, cluster significance threshold of *P<0.05*. Peak coordinates and cluster sizes are given.

**Table S6. Cluster values of the correlation between motor impairment and PPI**

| Correlation with motor impairment | x | y | z | Z | Cluster size |
|---|---|---|---|---|---|
| negative PPI IL SMA - spinal ~ NHP ratio | 5 | -4 | -173 | 4.14 | 460 |
| positive PPI IL SMA - spinal ~ UFMA | 4 | -44 | -173 | 4.16 | 154 |
| positive PPI IL PMV- spinal ~ NHP ratio | 1 | -49 | -167 | 4.5 | 1090 |
| negative PPI IL PMV - spinal ~ UFMA | 5 | -45 | -170 | 4.15 | 242 |

Stroke patients, threshold *Z>2.4*, cluster significance threshold of *P<0.05*. Peak coordinates and cluster sizes are given.
